# Noninvasive Hypokalemia Detection from Single-Lead AI-ECG: Development, Multicenter Validation, and Prospective Pilot Study in the Emergency Department

**DOI:** 10.64898/2026.05.23.26353774

**Authors:** Gongzheng Tang, Xiaojing Li, Yujie Xiao, Kexin Wang, Meng Xu, Xingliang Wu, Zheng Wei, Miao Yu, Xiaolan Chen, Wei Hong, Fang Cheng, Xiuqing Li, Jianxia Zhang, Shenda Hong

## Abstract

Hypokalemia is a common and potentially life-threatening electrolyte abnormality in emergency care, yet rapid noninvasive screening remains difficult in time-critical triage settings. We developed PocketED-K, a single-lead AI-ECG prescreening model initialized from ECGFounder, and evaluated it in retrospective multicenter cohorts and a prospective handheld pilot. Retrospective development and validation included 37,115 patients from MC-MED and MIMIC-ED, and the pilot enrolled 18 patients at Peking University First Hospital. Hypokalemia was defined as venous serum potassium < 3.5 mmol/L. PocketED-K achieved AUROCs of 0.8189 (95% CI 0.8172–0.8207) in internal testing, 0.8104 (95% CI 0.8092–0.8115) in temporal validation, and 0.7889 (95% CI 0.7692–0.8074) in independent external validation; external negative predictive value was 0.9911 (95% CI 0.9895–0.9925). Higher predicted risk was associated with ST-segment depression, T-wave flattening or inversion, and relative U-wave prominence. The prospective handheld pilot provided an initial signal of workflow feasibility in real-world acquisition. These findings support single-lead AI-ECG as a low-burden prescreening tool to prioritize potassium testing in emergency care.

## INTRODUCTION

Hypokalemia, defined as a venous serum potassium concentration below 3.5 mmol/L, is a frequent and clinically important electrolyte disturbance in emergency medicine^1,2^. Severe hypokalemia is associated with potentially life-threatening complications, including arrhythmias, paralysis, rhabdomyolysis, and diaphragmatic weakness^3–5^. Epidemiological studies have further suggested a nonlinear relationship between potassium homeostasis and mortality, with lower potassium levels associated with worse in-hospital outcomes^6,7^. These considerations underscore the clinical importance of early recognition, which remains challenging during rapid emergency assessment.

In routine care, hypokalemia is confirmed by venous potassium measurement, which is accurate but not immediately available at the moment when triage decisions are made. The electrocardiogram offers an appealing noninvasive alternative because potassium depletion can alter repolarization morphology, classically producing T-wave flattening, ST depression, and U-wave accentuation^8–10^. However, these changes are often subtle, variably expressed, and difficult to detect reliably by visual review alone. As a result, ECG has clinical relevance for hypokalemia, but has not been fully translated into a practical standalone prescreening tool.

Artificial intelligence, particularly deep learning, has been increasingly adopted in clinical medicine^11–17^. In electrocardiography, AI-based methods can detect subtle waveform patterns that are difficult to recognize by routine visual inspection and have shown promise for identifying cardiovascular, metabolic, and related abnormalities.^18–23^. For hypokalemia, 12-lead ECG models have demonstrated diagnostic potential, but the acquisition workflow of standard 12-lead ECG limits scalability for rapid prescreening and repeated measurement in busy emergency settings^24–26^. By contrast, single-lead ECG is inherently compatible with handheld devices and wearable platforms, making it more suitable for low-burden deployment. However, evidence for single-lead AI-based hypokalemia assessment remains limited, especially in terms of external validation across heterogeneous health systems and feasibility in prospective real-world use.

We therefore developed a single-lead AI-ECG model for hypokalemia prescreening in the emergency department, initialized from the pretrained ECGFounder foundation model, and evaluated it across three retrospective validation settings: an internal test set, a temporal validation set, and an independent external validation set. To further examine translational potential, we also conducted a prospective handheld pilot study focused on real-world workflow feasibility.

## RESULTS

### Study Overview

We developed PocketED-K by fine-tuning a pretrained ECG foundation model on paired lead I ECG and serum potassium data (Figure 1). In MC-MED, patients were chronologically partitioned according to the timestamp of their first eligible ECG–potassium pair. Patients whose first eligible pair occurred before the predefined temporal cutoff constituted the development cohort, which was further split at the patient level into fine-tuning, model-selection, and internal test subsets. Patients whose first eligible pair occurred after the cutoff constituted the temporal validation cohort. To minimize patient-level information leakage, any patient included in the development cohort was excluded from temporal validation even if later eligible records were available; therefore, no patient appeared in more than one MC-MED retrospective partition. MIMIC-ED was reserved exclusively for independent external validation. A separate prospective pilot at the Emergency Department of Peking University First Hospital evaluated the feasibility of a smartphone-connected handheld acquisition and inference workflow and was not used to estimate definitive clinical performance.

**Figure 1:**
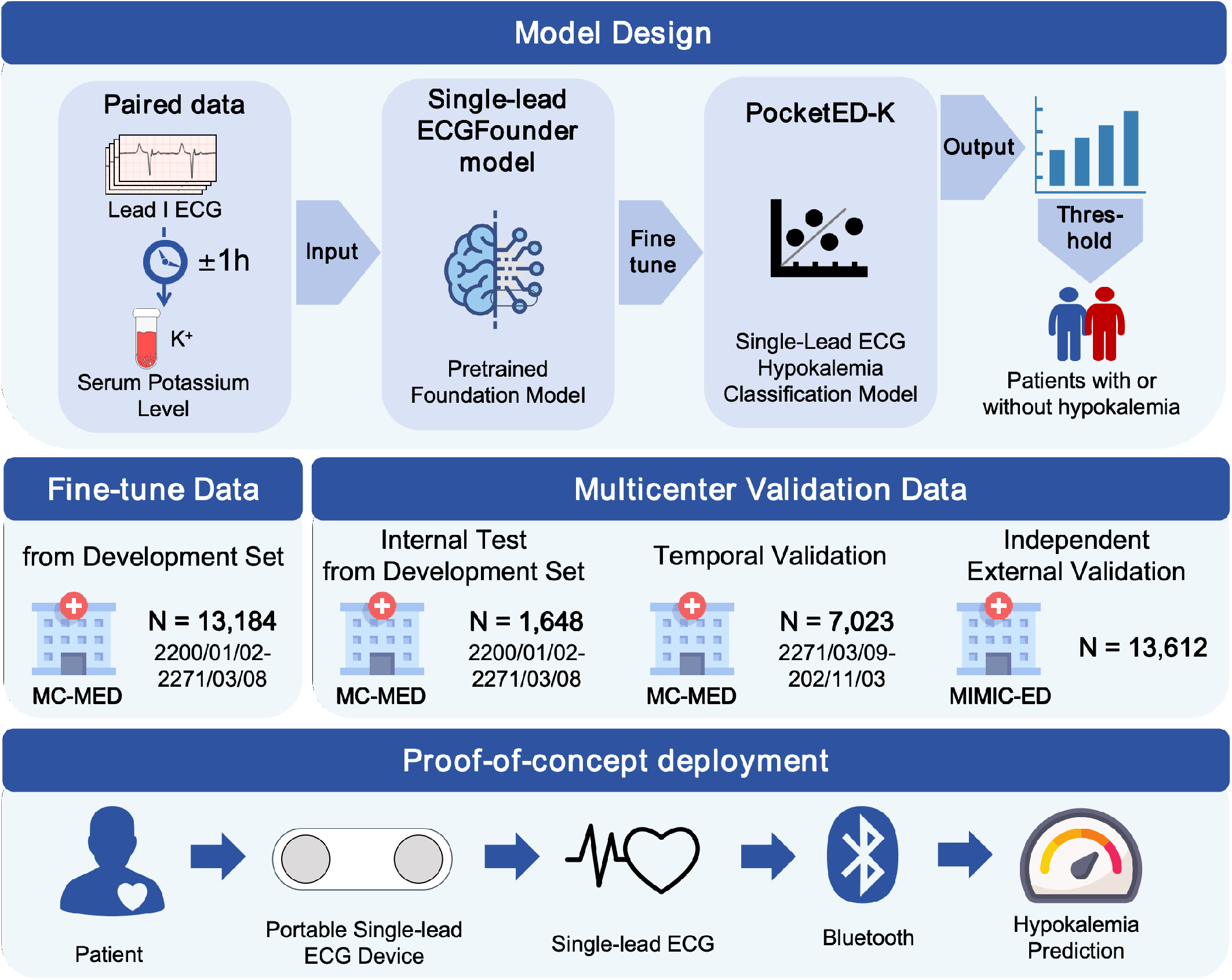
Overview of model design, multicenter retrospective validation, and prospective pilot deployment of PocketED-K. Paired data were constructed by linking lead I ECG recordings to serum potassium measurements obtained within a *±* 1-hour window. These paired ECG–*K*^+^ data were used to fine-tune a single-lead ECGFounder foundation model to develop PocketED-K for hypokalemia classification. In MC-MED, the development set provided a fine-tuning subset, a model-selection subset, and an internal test subset, whereas a temporally later subset was used for temporal validation. Independent external validation was performed in MIMIC-ED. A proof-of-concept handheld deployment workflow was also implemented, in which a portable single-lead ECG device acquired ECG signals and transmitted them through Bluetooth to support hypokalemia prediction in a smartphone-connected pipeline.

### Study population and baseline characteristics

The retrospective study population included 37,115 unique patients from MC-MED and MIMIC-ED, comprising 16,480 patients in the MC-MED development set, 7,023 patients in the MC-MED temporal validation set, and 13,612 patients in the MIMIC-ED independent external validation set. The prospective in-hospital pilot evaluation of the handheld workflow additionally included 18 patients with 32 handheld recordings at Peking University First Hospital and was analysed separately from the retrospective cohorts.

Baseline demographic and clinical characteristics, laboratory measurements, ECG–potassium pairing intervals, and comorbidities across the retrospective study cohorts are summarized in Table 1. Across the three retrospective cohorts, mean age ranged from 59.87 to 63.61 years, and men accounted for 50.3%–52.7% of patients. Compared with the MC-MED development and temporal validation sets, the independent external validation cohort from MIMIC-ED was older and had a substantially greater burden of renal dysfunction and cardiorenal comorbidity, reflected by higher serum creatinine, lower eGFR, and higher prevalences of chronic kidney disease, heart failure, and coronary artery disease.

**Table 1:**
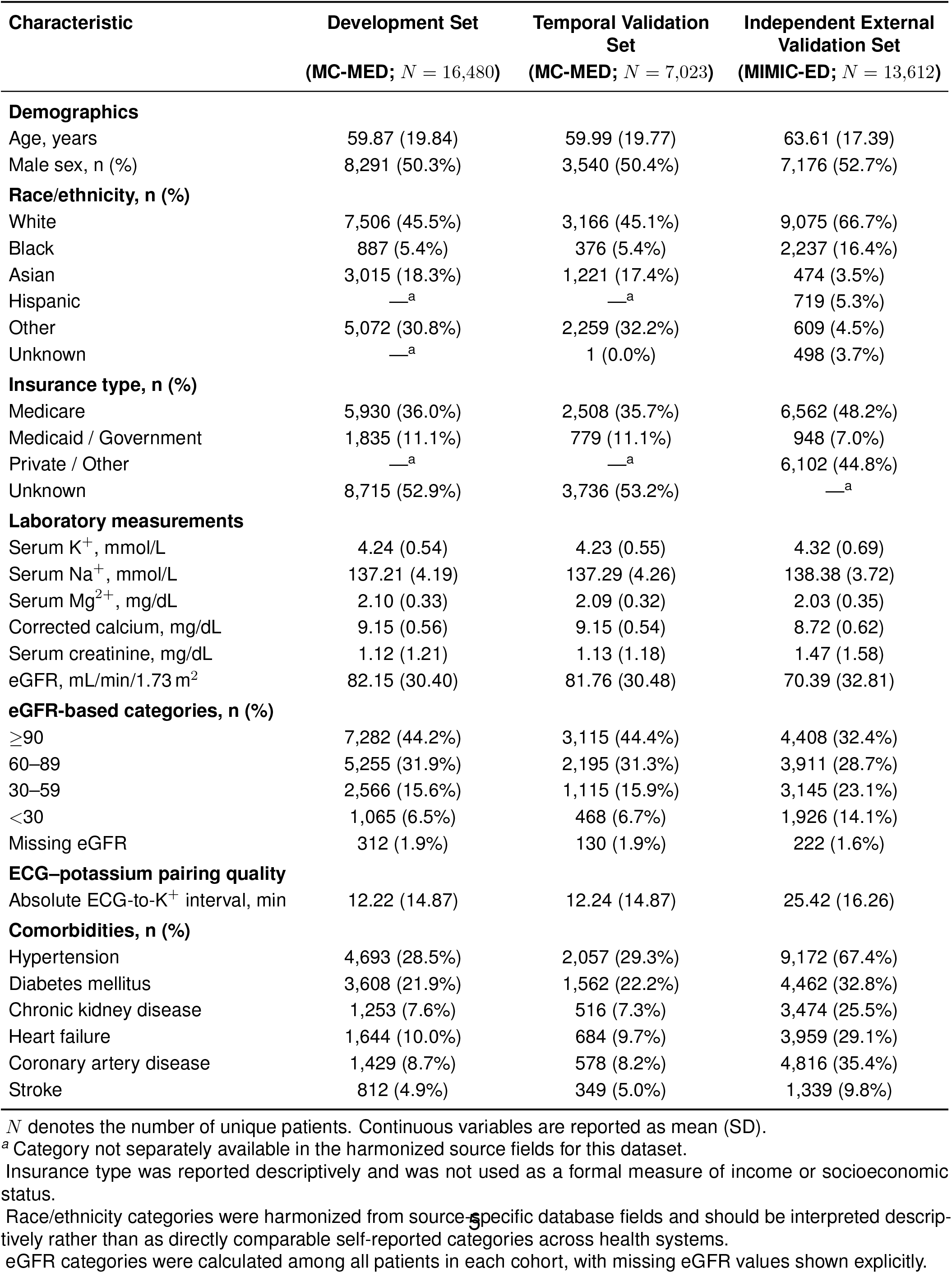
Baseline demographic and clinical characteristics, laboratory measurements, ECG– potassium pairing intervals, and comorbidities across the study cohorts. Continuous variables are presented as mean (SD), and categorical variables as n (%). *K*^+^: serum potassium; eGFR: estimated glomerular filtration rate; ECG: electrocardiogram; MC-MED: Multimodal Clinical Monitoring in the Emergency Department; MIMIC-ED: Medical Information Mart for Intensive Care-Emergency Department.

| Characteristic | Development Set<br>(MC-MED; $N = 16,480$) | Temporal Validation Set<br>(MC-MED; $N = 7,023$) | Independent External Validation Set<br>(MIMIC-ED; $N = 13,612$) |
| --- | --- | --- | --- |
| **Demographics** |  |  |  |
| Age, years | 59.87 (19.84) | 59.99 (19.77) | 63.61 (17.39) |
| Male sex, n (%) | 8,291 (50.3%) | 3,540 (50.4%) | 7,176 (52.7%) |
| **Race/ethnicity, n (%)** |  |  |  |
| White | 7,506 (45.5%) | 3,166 (45.1%) | 9,075 (66.7%) |
| Black | 887 (5.4%) | 376 (5.4%) | 2,237 (16.4%) |
| Asian | 3,015 (18.3%) | 1,221 (17.4%) | 474 (3.5%) |
| Hispanic | —<sup>a</sup> | —<sup>a</sup> | 719 (5.3%) |
| Other | 5,072 (30.8%) | 2,259 (32.2%) | 609 (4.5%) |
| Unknown | —<sup>a</sup> | 1 (0.0%) | 498 (3.7%) |
| **Insurance type, n (%)** |  |  |  |
| Medicare | 5,930 (36.0%) | 2,508 (35.7%) | 6,562 (48.2%) |
| Medicaid / Government | 1,835 (11.1%) | 779 (11.1%) | 948 (7.0%) |
| Private / Other | —<sup>a</sup> | —<sup>a</sup> | 6,102 (44.8%) |
| Unknown | 8,715 (52.9%) | 3,736 (53.2%) | —<sup>a</sup> |
| **Laboratory measurements** |  |  |  |
| Serum $K^+$, mmol/L | 4.24 (0.54) | 4.23 (0.55) | 4.32 (0.69) |
| Serum $Na^+$, mmol/L | 137.21 (4.19) | 137.29 (4.26) | 138.38 (3.72) |
| Serum $Mg^{2+}$, mg/dL | 2.10 (0.33) | 2.09 (0.32) | 2.03 (0.35) |
| Corrected calcium, mg/dL | 9.15 (0.56) | 9.15 (0.54) | 8.72 (0.62) |
| Serum creatinine, mg/dL | 1.12 (1.21) | 1.13 (1.18) | 1.47 (1.58) |
| eGFR, mL/min/1.73 m<sup>2</sup> | 82.15 (30.40) | 81.76 (30.48) | 70.39 (32.81) |
| **eGFR-based categories, n (%)** |  |  |  |
| ≥90 | 7,282 (44.2%) | 3,115 (44.4%) | 4,408 (32.4%) |
| 60–89 | 5,255 (31.9%) | 2,195 (31.3%) | 3,911 (28.7%) |
| 30–59 | 2,566 (15.6%) | 1,115 (15.9%) | 3,145 (23.1%) |
| <30 | 1,065 (6.5%) | 468 (6.7%) | 1,926 (14.1%) |
| Missing eGFR | 312 (1.9%) | 130 (1.9%) | 222 (1.6%) |
| **ECG–potassium pairing quality** |  |  |  |
| Absolute ECG-to-$K^+$ interval, min | 12.22 (14.87) | 12.24 (14.87) | 25.42 (16.26) |
| **Comorbidities, n (%)** |  |  |  |
| Hypertension | 4,693 (28.5%) | 2,057 (29.3%) | 9,172 (67.4%) |
| Diabetes mellitus | 3,608 (21.9%) | 1,562 (22.2%) | 4,462 (32.8%) |
| Chronic kidney disease | 1,253 (7.6%) | 516 (7.3%) | 3,474 (25.5%) |
| Heart failure | 1,644 (10.0%) | 684 (9.7%) | 3,959 (29.1%) |
| Coronary artery disease | 1,429 (8.7%) | 578 (8.2%) | 4,816 (35.4%) |
| Stroke | 812 (4.9%) | 349 (5.0%) | 1,339 (9.8%) |

$N$ denotes the number of unique patients. Continuous variables are reported as mean (SD).
<sup>a</sup> Category not separately available in the harmonized source fields for this dataset.
Insurance type was reported descriptively and was not used as a formal measure of income or socioeconomic status.
Race/ethnicity categories were harmonized from source-specific database fields and should be interpreted descriptively rather than as directly comparable self-reported categories across health systems.
eGFR categories were calculated among all patients in each cohort, with missing eGFR values shown explicitly.

The mean absolute ECG-to-*K*^+^ interval was 12.22 min in the development set, 12.24 min in the temporal validation set, and 25.42 min in the independent external validation set, indicating modest variation in pairing latency across cohorts while remaining within a clinically relevant time window.

### PocketED-K generalized across internal, temporal, and independent external validation sets

PocketED-K showed stable discrimination across the validation settings. In the internal test set, the AUROC was 0.8189 (95% CI 0.8172–0.8207), with sensitivity of 0.7837 (95% CI 0.7802– 0.7871), specificity of 0.6989 (95% CI 0.6980–0.6997), and negative predictive value of 0.9858 (95% CI 0.9855–0.9860). In the temporal validation set, the AUROC was 0.8104 (95% CI 0.8092–0.8115), with sensitivity of 0.7318 (95% CI 0.7294–0.7343), specificity of 0.7364 (95% CI 0.7359–0.7370), and negative predictive value of 0.9816 (95% CI 0.9814–0.9818), indicating robustness to temporal drift. In the independent external validation set, the AUROC was 0.7889 (95% CI 0.7692–0.8074), with sensitivity of 0.7388 (95% CI 0.6981–0.7757), specificity of 0.7094 (95% CI 0.7031–0.7156), and negative predictive value of 0.9911 (95% CI 0.9895–0.9925), despite differences in ECG platform and disease prevalence, as shown in Figure 2.

**Figure 2:**
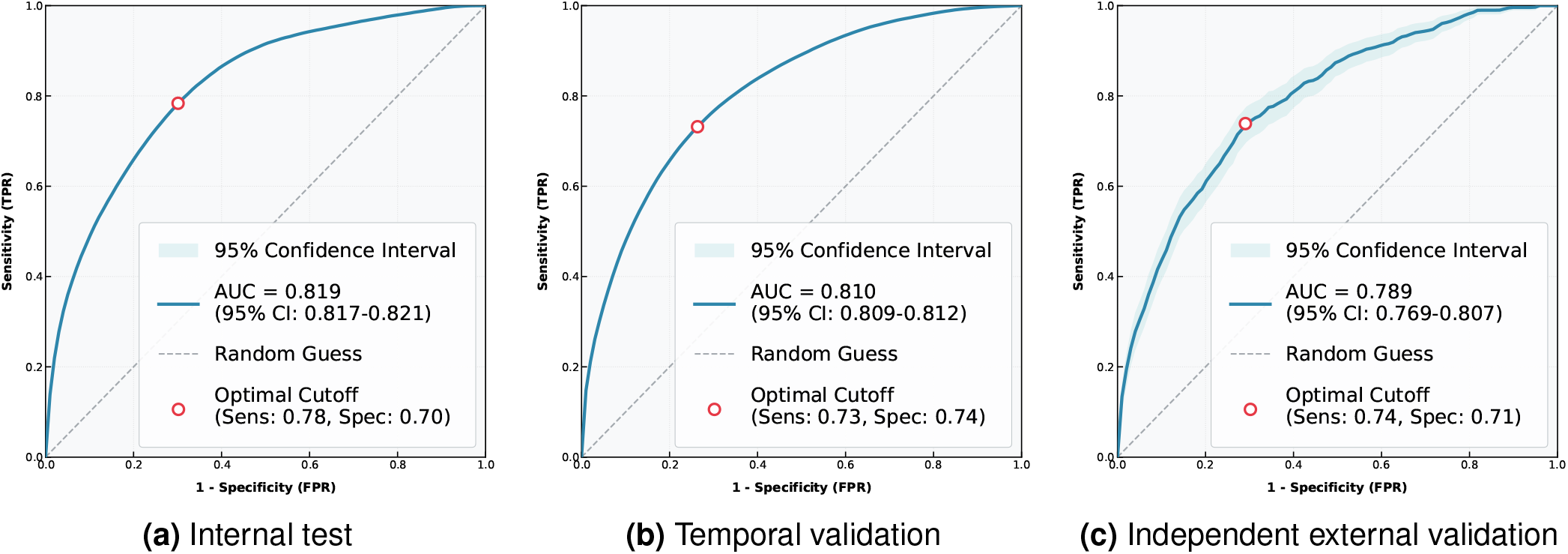
ROC curves for evaluation of model performance. (a) ROC curve for internal testing in MC-MED. (b) ROC curve for temporal validation in MC-MED. (c) ROC curve for independent external validation in MIMIC-ED.

Although discrimination decreased across increasingly heterogeneous evaluation settings, negative predictive value remained high, reaching 0.9911 (95% CI 0.9895–0.9925) in the independent external validation set. By contrast, positive predictive value was modest across datasets, which indicates that PocketED-K is best positioned as a rule-out prescreening tool rather than a replacement for laboratory confirmation.

Predicted hypokalemia risk increased progressively across serum potassium intervals in all three cohorts (Figure 3). Samples with lower potassium concentrations were assigned systematically higher model outputs, whereas most normokalemic samples clustered at low predicted risk. This pattern was preserved in internal testing, temporal validation, and independent external validation, indicating that model output retained a clinically coherent gradient across datasets despite distributional shift.

**Figure 3:**
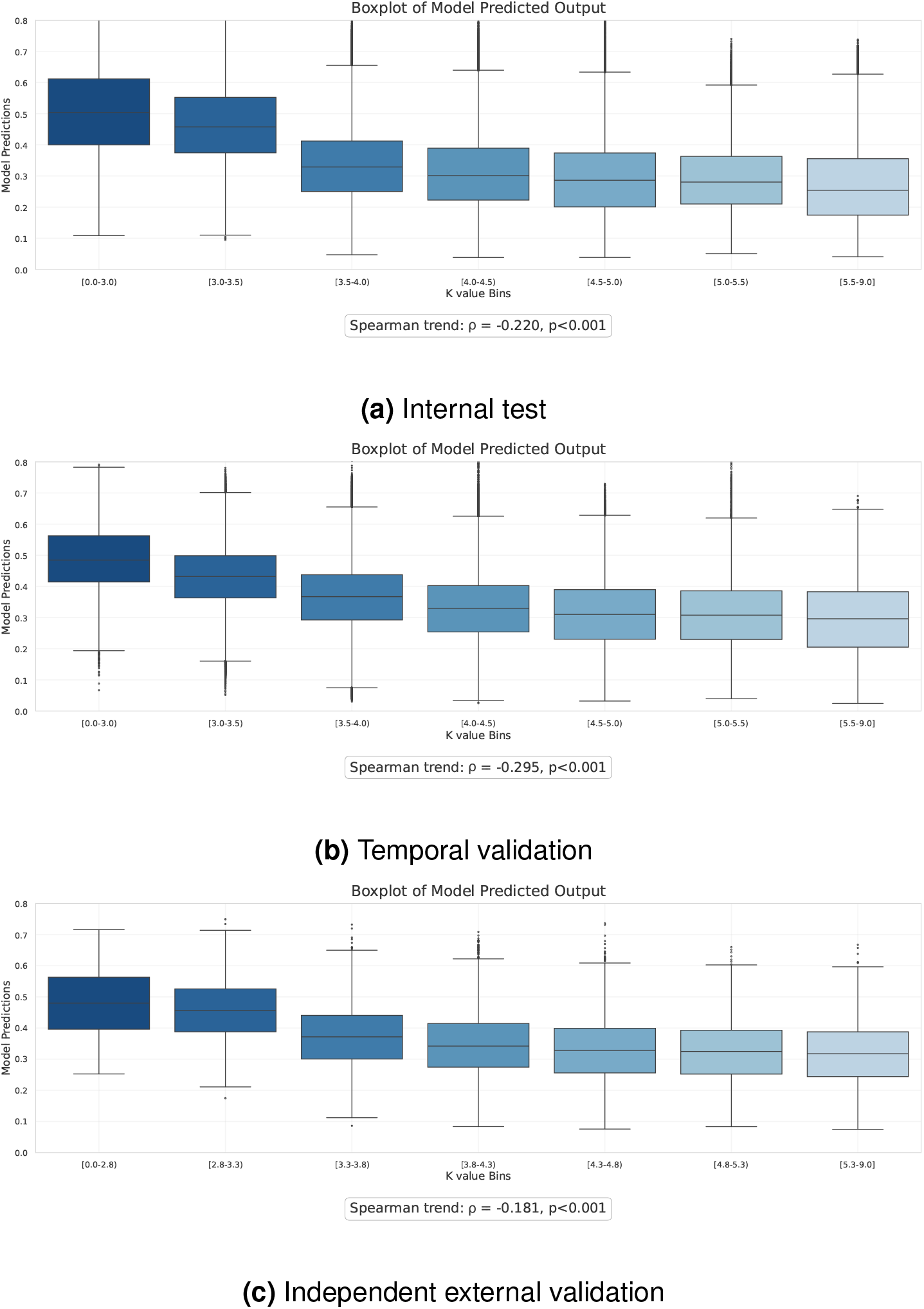
Boxplots of model-predicted hypokalemia risk across serum potassium intervals. Predicted risk is shown across dataset-specific serum potassium bins. For the MC-MED internal test and temporal validation sets, serum potassium was grouped into 0–3.0, 3.0–3.5, 3.5–4.0, 4.0–4.5, 4.5–5.0, 5.0–5.5, and 5.5–9.0 mmol/L. For the MIMIC-ED independent external validation set, serum potassium was grouped into 0–2.8, 2.8–3.3, 3.3–3.8, 3.8–4.3, 4.3–4.8, 4.8–5.3, and 5.3–9.0 mmol/L, reflecting the local reference range in MIMIC-ED. Each box shows the median and interquartile range, whiskers extend to 1.5 *×* IQR, and points indicate outliers. (a) Internal test set from MC-MED. (b) Temporal validation set from MC-MED. (c) Independent external validation set from MIMIC-ED.

### PocketED-K captured recognizable electrophysiological features of hypokalemia

Signal-averaged lead I waveforms differed clearly between low-risk and high-risk groups (Figure 4). Relative to the low-risk group, the high-risk group showed ST-segment depression, T-wave flattening or inversion, and relative U-wave prominence; a modest increase in P-wave amplitude was also observed. These patterns are consistent with established electrophysiological manifestations of hypokalemia and support the physiological plausibility of the model output.

**Figure 4:**
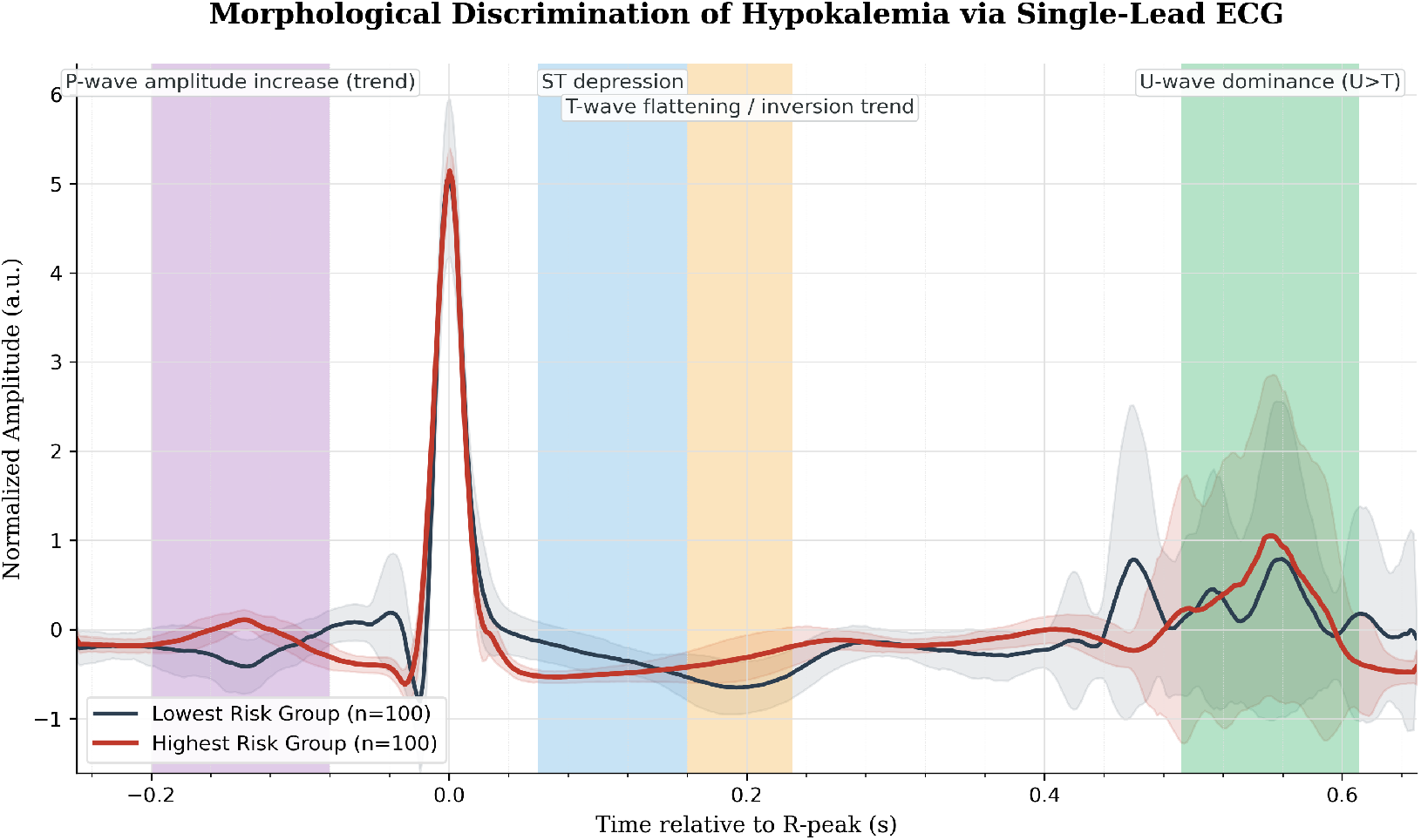
Signal-averaged waveform comparison between the lowest-risk and highest-risk groups. Average lead I ECG waveforms are shown for the lowest-risk group (blue) and the highest-risk group (red), defined according to model-predicted hypokalemia risk. Shaded areas indicate standard deviation. The highest-risk group showed recognizable morphological changes consistent with hypokalemia, including ST-segment depression, T-wave flattening or inversion, and relative U-wave prominence.

**Figure 5:**
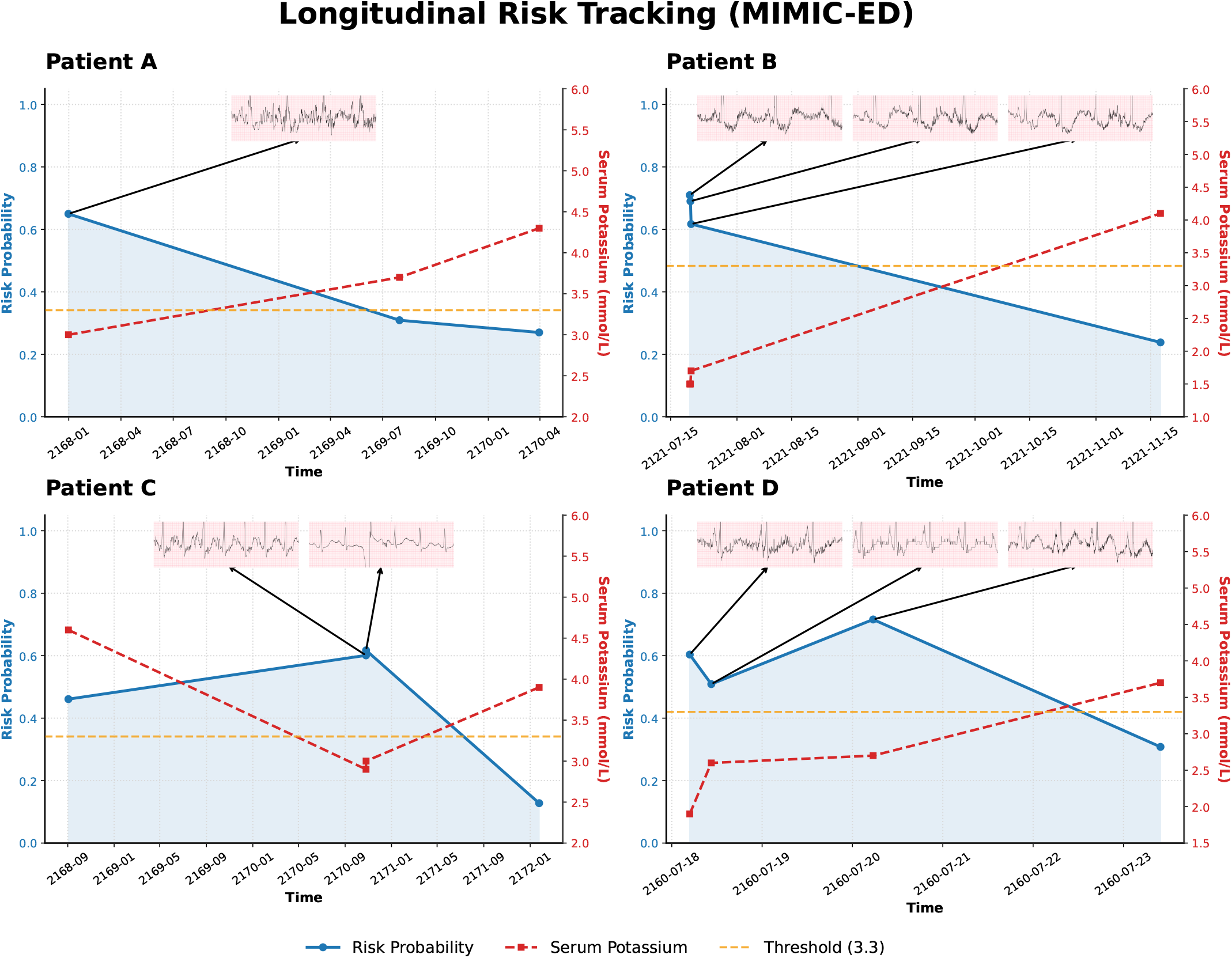
Longitudinal tracking of serum potassium and model-predicted hypokalemia risk in representative de-identified participants from the independent external validation cohort, MIMIC-ED. The red line indicates laboratory serum potassium, and the blue dashed line indicates model-predicted risk of hypokalemia. In this analysis, low-potassium status was defined as serum *K*^+^ *<* 3.3 mmol/L, consistent with the reference range used in MIMIC-ED. Repeated measurements are shown according to within-participant temporal order rather than exact calendar dates. Across all four cases, model-predicted risk increased during low-potassium episodes and decreased after potassium recovery, indicating that PocketED-K tracked clinically meaningful within-participant potassium dynamics over time.

### Longitudinal case studies supported within-participant tracking of hypokalemia risk

Longitudinal case studies were performed in four de-identified participants from the independent external validation cohort, MIMIC-ED, who had repeated paired ECG–*K*^+^ measurements over time. Because the normal potassium reference range in MIMIC-ED was defined as 3.3–5.3 mmol/L, low-potassium status in this analysis was defined as serum *K*^+^ *<* 3.3 mmol/L. To minimize the risk of participant identification, individual-level demographic details, specific diagnoses, and exact calendar dates were not reported; repeated measurements are presented only according to within-participant temporal order.

Across the four participants, model-predicted risk was consistently higher at low-potassium timepoints than at non-low-potassium timepoints. Participant A contributed three paired ECG– *K*^+^ timepoints, with serum potassium ranging from 3.0 to 4.3 mmol/L and model-predicted risk ranging from 0.270 to 0.650. Risk was highest at the low-potassium timepoint (*K*^+^=3.0 mmol/L; risk, 0.650), whereas the mean risk at non-low-potassium timepoints was 0.290, yielding a within-participant contrast of 0.360.

Participant B contributed four paired timepoints, with potassium values ranging from 1.5 to 4.1 mmol/L and predicted risk ranging from 0.238 to 0.710. Three consecutive severe low-potassium measurements (*K*^+^=1.5, 1.5, and 1.7 mmol/L) were associated with persistently high predicted risks of 0.710, 0.690, and 0.617, respectively, whereas risk decreased to 0.238 after potassium returned to 4.1 mmol/L. The mean risk difference between low-potassium and non-low-potassium states was 0.434.

Participant C contributed four paired timepoints, with potassium ranging from 2.9 to 4.6 mmol/L and predicted risk ranging from 0.128 to 0.617. During two low-potassium episodes (*K*^+^=2.9 and 3.0 mmol/L), predicted risks were 0.601 and 0.617, compared with 0.461 and 0.128 at non-low-potassium timepoints, yielding a within-participant contrast of 0.315.

Participant D contributed four paired timepoints, with potassium ranging from 1.9 to 3.7 mmol/L and predicted risk ranging from 0.308 to 0.716. Elevated risks were observed throughout the low-potassium phase, with predicted risks of 0.604, 0.509, and 0.716 at *K*^+^=1.9, 2.6, and 2.7 mmol/L, respectively, followed by a decline to 0.308 after recovery to *K*^+^=3.7 mmol/L. The mean risk difference between low-potassium and non-low-potassium states was 0.301.

Overall, these longitudinal case studies show that PocketED-K risk estimates varied concordantly with serum potassium status within the same participant across repeated measurements. These findings support the potential utility of PocketED-K for repeated prescreening and dynamic monitoring in external real-world settings, while avoiding disclosure of participant-level identifying information.

### PocketED-K supported near-real-time inference in a handheld workflow

In a prospective pilot at Peking University First Hospital, 18 patients contributed 32 handheld recordings. PocketED-K generated risk estimates during routine acquisition, and the single low-potassium case in the pilot cohort was correctly flagged (serum potassium 3.4 mmol/L; Fig. 6). Given the small sample size and single event, these data should be interpreted as preliminary evidence of workflow feasibility rather than a definitive estimate of clinical performance.

**Figure 6:**
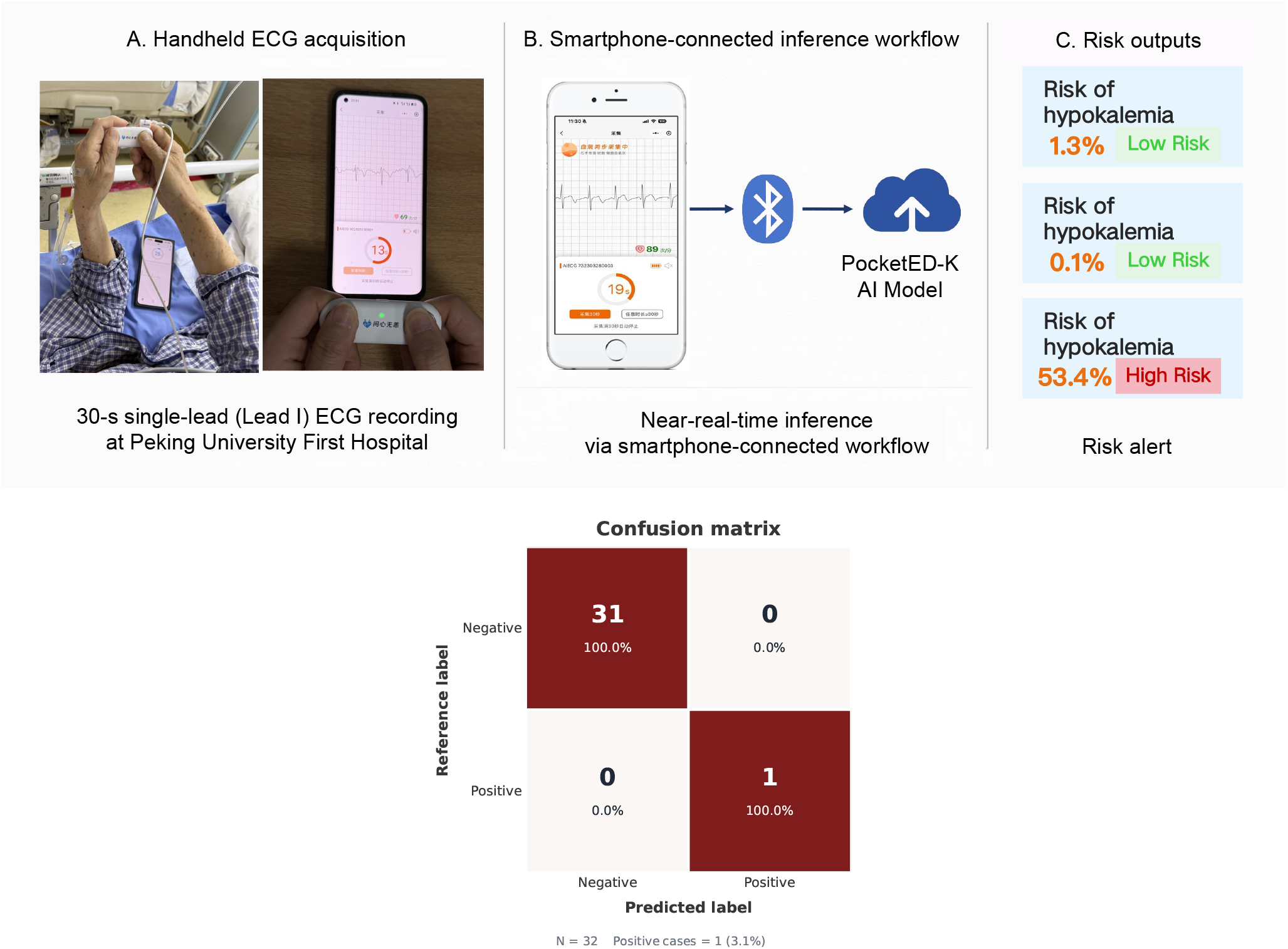
Handheld implementation and prospective pilot screening of PocketED-K. The top panel shows the proof-of-concept handheld workflow, in which a portable single-lead ECG device paired with a smartphone application was used to acquire 30-s lead I recordings at Peking University First Hospital, followed by smartphone-assisted transmission and PocketED-K analysis for near-real-time hypokalemia risk alerting. The bottom panel shows the confusion matrix of the prospective pilot screening cohort, comprising 18 patients and 32 handheld recordings.

## DISCUSSION

This study adds to the emerging literature on AI-enabled electrolyte prescreening by showing that a foundation-model-initialized single-lead AI-ECG approach can identify hypokalemia with clinically meaningful discrimination across temporally separated and externally independent retrospective validation settings. The main value of PocketED-K is not that it replaces laboratory testing, but that it may support earlier risk stratification in situations in which blood testing is delayed, repeated surveillance is needed, or ECG acquisition is more accessible than immediate biochemical assessment.

Existing literature on AI-ECG for electrolyte disorders has been dominated by studies based on standard 12-lead ECG, which generally report stronger discrimination than single-lead approaches and have established proof of principle that deep-learning models can detect potassium-related ECG signatures beyond routine visual interpretation. Evidence for single-lead hypokalemia detection remains comparatively limited and has largely been derived from smaller studies with narrower populations, less rigorous transport testing, or limited assessment across acquisition settings. Against this background, the present study extends prior work by evaluating a single-lead model in a multicenter setting with temporal separation within one health system, independent external validation in another health system, waveform-level interpretability analysis, longitudinal case review, and a prospective pilot handheld workflow.

The pattern of performance observed in this study is clinically informative. Although discrimination declined from internal testing to temporal and external validation, negative predictive value remained high across cohorts, particularly in external validation. This pattern is consistent with the expected behavior of a rule-out prescreening tool under prevalence shift and supports a clinically plausible use case. In emergency care, a model of this type may be most useful for identifying patients at sufficiently low probability of hypokalemia to defer urgent repeat testing, while prioritizing confirmatory laboratory assessment in those with elevated predicted risk.

The biological plausibility of the model is supported by the waveform analyses. Signal-averaged comparisons showed that higher model-predicted risk was associated with U-wave prominence, T-wave flattening or inversion, and ST-segment depression, all of which are well-recognized electrophysiological manifestations of hypokalemia. This observation is relevant because one of the recurring concerns in AI-ECG research is whether model outputs reflect meaningful physiology or spurious correlates. Although group-level morphology analysis cannot fully resolve that question, the concordance between known hypokalemia-related features and the high-risk waveform pattern supports the view that the model is leveraging clinically interpretable signal structure rather than only hidden dataset artifacts.

The longitudinal case studies provide a complementary perspective that is often missing from cross-sectional model evaluations. In the external validation cohort, model-predicted risk changed in parallel with within-patient potassium trajectories across repeated measurements, even though the selected cases varied in age, comorbidity burden, and diagnosis context. This does not establish that the model can replace serial laboratory testing, but it does suggest that the output of PocketED-K may track evolving potassium-related electrophysiology over time. From a translational standpoint, this feature is potentially important, because repeated monitoring is one of the clinical scenarios in which low-burden single-lead systems may offer the greatest practical value.

The handheld implementation further strengthens the translational relevance of the study, although the current evidence should be interpreted cautiously. Prior work in digital cardiology has shown that technical feasibility alone does not guarantee reliable real-world performance once models are deployed on native handheld or wearable recordings, where motion artifact, contact instability, user variability, and altered preprocessing pipelines can all affect signal quality and calibration. The present prototype therefore serves primarily as a proof of workflow feasibility. The preliminary in-hospital evaluation at Peking University First Hospital suggests that near-real-time inference can be integrated into routine acquisition and that clinically relevant low-potassium events can be flagged in practice, but these early observations should not be overinterpreted as evidence of clinical effectiveness.

Several broader implications follow from these findings. First, the study supports the idea that foundation-model initialization may be useful even in relatively signal-limited single-lead tasks, particularly when the target phenotype is subtle and training labels are derived from routine care. Second, the results highlight the importance of testing transportability across health systems and acquisition environments. The performance gap between internal and external evaluation was not a weakness to be explained away, but rather an expected and informative feature of clinically realistic validation. Third, the combination of high negative predictive value, waveform plausibility, and dynamic within-patient tracking suggests that the most appropriate future applications of this class of model may lie in prescreening, serial surveillance, and escalation support, especially in care pathways in which rapid blood testing is not always immediately available.

This study has several limitations. First, the main analyses were retrospective and based on routinely collected clinical data, which limits causal inference regarding the effect of AI-guided screening on workflow or patient outcomes. Second, although temporal and independent external validation were included, additional validation across broader geographic, ethnic, and device-level settings remains necessary. Third, the single-lead input was derived primarily from clinical ECG systems rather than from native consumer-grade recordings, and model calibration may differ when signal characteristics change. Finally, the preliminary handheld evaluation was small and should be regarded as an initial translational signal rather than a definitive assessment of deployment readiness.

In conclusion, PocketED-K provides multicenter evidence that a foundation-model-initialized single-lead AI-ECG approach can support hypokalemia prescreening across temporally separated and externally independent retrospective validation settings. Rather than serving as a replacement for laboratory testing, its most plausible clinical role is as a scalable rule-out and surveillance-support tool. The prospective handheld pilot further provided an initial signal of workflow feasibility. A multicenter prospective study in native handheld and wearable environments is planned as the next step to evaluate calibration, usability, and clinical effectiveness.

## METHODS

### Study design and data sources

We conducted a multicohort study using de-identified ECG and laboratory records collected during routine clinical care, together with a separate prospective pilot of handheld ECG acquisition. Reporting followed TRIPOD+AI recommendations^27^. The retrospective analyses used two data sources from independent health systems. The primary development and temporal validation data were derived from Multimodal Clinical Monitoring in the Emergency Department (MC-MED), which includes monitored-bed emergency department visits at Stanford Health Care between 2020 and 2022. For independent external validation, we used the full MIMIC-IV database records from Beth Israel Deaconess Medical Center between 2011 and 2019 and constructed an emergency department cohort by restricting the source population to eligible ED encounters. In this study, this MIMIC-IV-derived emergency department cohort is referred to as MIMIC-ED for brevity. MC-MED served as the primary dataset for model development and temporal validation, whereas MIMIC-ED was used exclusively for independent external validation. To explore workflow feasibility outside the retrospective datasets, we additionally performed a prospective in-hospital pilot of handheld lead-I acquisition at the Emergency Department of Peking University First Hospital. Ethical approval was obtained from the Biomedical Ethics Committee of Peking University First Hospital (2024yan655-002). The requirement for informed consent was waived for the retrospective analyses because only de-identified routinely collected data were used. Written informed consent was obtained from participants in the prospective handheld pilot before ECG acquisition.

### Study population assembly and data partitioning

As illustrated in Figure 7, we first screened all patients with available ECG data during the study periods. After exclusion of haemolysed samples, venous serum potassium measurements were linked to ECG recordings using an ECG-anchored pairing strategy. Specifically, for each ECG, we searched for potassium measurements obtained within a *±* 1-hour window and retained the closest eligible measurement to form a unique ECG–potassium pair. ECGs without any eligible potassium measurement within the predefined time window were excluded.

**Figure 7:**
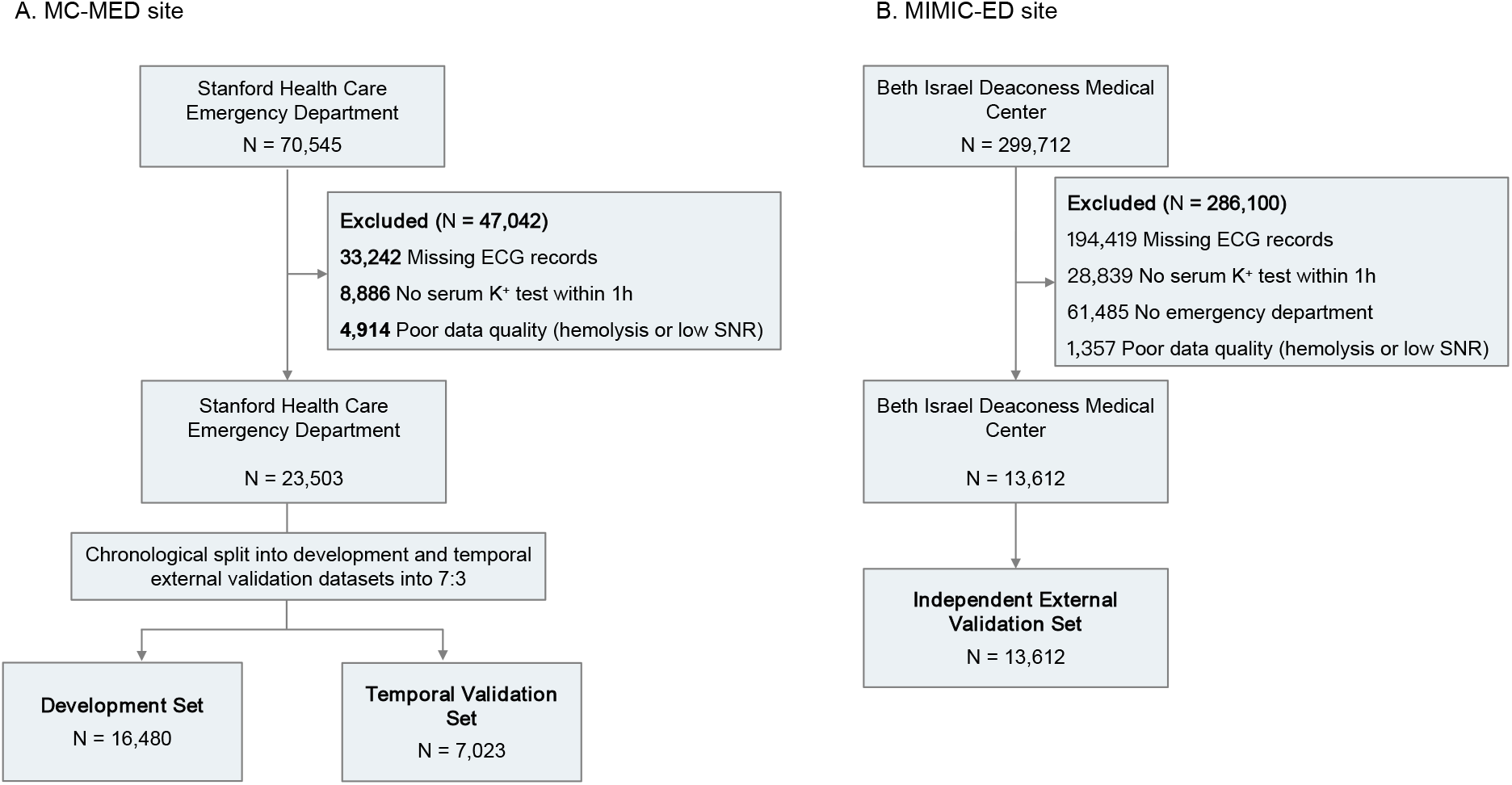
STARD flowchart for study population selection and dataset partitioning. Panel **A** shows cohort construction in the primary dataset, MC-MED, derived from Stanford Health Care Emergency Department. After exclusion of records with missing ECG data, no eligible serum *K*^+^ measurement within 1 hour, or poor data quality, the remaining patients were ordered chronologically and divided into development and temporal validation datasets in a 7:3 ratio. Panel **B** shows cohort construction for the independent external validation cohort derived from the full MIMIC-IV v2.2 database at Beth Israel Deaconess Medical Center. After restriction to emergency department encounters, records with missing ECG data, no eligible serum *K*^+^ measurement within 1 hour, or poor data quality were excluded. The remaining patients constituted the MIMIC-IV-derived ED external validation cohort. In all panels, *N* denotes the number of unique patients.

In MC-MED, patients were first ordered chronologically and then divided in a 7:3 ratio. The earlier 70% constituted the development set, within which patients were further divided at the patient level into a fine-tuning subset, a model-selection subset, and an internal test subset in an 8:1:1 ratio. The later 30% served as the temporal validation set. To reduce information leakage, any patient appearing in the development set was excluded from the temporal validation set, even if later records were available. MIMIC-ED was reserved exclusively for independent external validation.

Because individual patients could contribute multiple ECG–potassium pairs over time, the number of unique patients differed from the number of paired records. The prospective handheld pilot cohort at Peking University First Hospital was assembled separately and was not included in the retrospective development or validation partitions.

### Reference standard and single-lead signal preprocessing

The biochemical target for supervision was venous serum potassium. To maximize temporal consistency between electrophysiological input and laboratory reference, labeling was performed by assigning each ECG the nearest eligible potassium measurement within 1 hour after exclusion of haemolysed samples. Lead-I signals were then used as the sole model input. Before model ingestion, all recordings underwent the same preprocessing procedure: baseline drift and highfrequency artefacts were attenuated with a 0.5–40 Hz band-pass filter, continuous recordings were divided into non-overlapping 10-s segments, each segment was resampled to 500 Hz to conform to the foundation-model input format, and amplitude values were standardized by z-score normalization.

### Model development

PocketED-K was obtained by adapting ECGFounder to the hypokalemia classification task. ECGFounder is a large pretrained ECG representation model with a Net1D-derived backbone and stage-wise scaling strategy related to RegNet, trained on more than ten million recordings from multinational sources^28–30^. In the present work, pretrained weights were used as initialization for downstream fine-tuning on single-lead data. Optimization used binary cross-entropy as the objective function and Adam as the optimizer. Training proceeded for a maximum of 30 epochs, while AUROC on the model-selection subset was used to guide checkpoint selection. When validation performance plateaued for 10 successive epochs, the learning rate was decayed by a factor of 10. The checkpoint achieving the highest validation AUROC was carried forward for all subsequent evaluations.

### Performance evaluation and statistical analyses

The main endpoint was the ability to distinguish hypokalemia, defined as serum potassium below 3.5 mmol/L, from non-hypokalemic states. Discriminative performance was summarized primarily by AUROC. Threshold-dependent operating characteristics, including sensitivity, specificity, and negative predictive value, were reported at cohort-specific operating points derived from the corresponding ROC curves to describe the trade-off between sensitivity and specificity within each evaluation setting. Because multiple paired observations could originate from the same patient, confidence intervals were derived using patient-clustered bootstrap resampling with 2000 iterations rather than treating all pairs as independent. The prospective handheld pilot was analyzed separately and interpreted as a feasibility-oriented assessment rather than as a definitive estimate of clinical screening accuracy.

### Additional analyses and explainability

We performed additional analyses to evaluate clinically relevant model behaviour beyond binary discrimination. First, waveform-level explainability analysis was performed by stratifying samples according to model-predicted risk and comparing signal-averaged heartbeat morphology between high-risk and low-risk groups, with particular attention to atrial and repolarization phases. Second, we examined the distribution of model-predicted risk across serum potassium intervals in each cohort to assess whether model output preserved clinically coherent gradients across datasets.

To assess whether model outputs tracked disease dynamics at the individual level, we examined representative longitudinal patients with repeated ECG–potassium pairs over time in the independent external validation cohort. Because the normal potassium reference range in MIMIC-ED was defined as 3.3–5.3 mmol/L, low-potassium status in the case-study analysis was defined as serum *K*^+^ *<* 3.3 mmol/L. Dates in MIMIC-ED were shifted as part of the de-identification process and therefore preserved within-patient temporal order without corresponding to real calendar dates.

### Proof-of-concept handheld deployment

To explore real-world implementation, we established a prototype handheld workflow based on 30-s single-lead acquisition. Each handheld recording was partitioned into three contiguous 10-s segments so that inference matched the temporal granularity used during model training. Segment-level outputs were combined to produce a single recording-level risk estimate. Feasibility was examined in a preliminary in-hospital pilot at the Emergency Department of Peking University First Hospital, where 18 patients contributed 32 handheld recordings. The system generated risk predictions during routine acquisition and identified the only low-potassium event in the pilot cohort (serum potassium 3.4 mmol/L).

## Declaration statements

### Data Availability

The MC-MED data used for model development, internal testing, and temporal validation were obtained from PhysioNet and are available to qualified researchers under credentialed access at https://physionet.org/content/mc-med/1.0.1/. The MIMIC-IV-derived emergency-department data used for independent external validation were obtained from PhysioNet under credentialed access, including MIMIC-IV clinical records at https://physionet.org/content/mimiciv/2.2/ and MIMIC-IV-ECG records at https://physionet.org/content/mimic-iv-ecg/1.0/. Access to these PhysioNet resources is available to researchers who complete the required training, approval process, and data-use agreement.

The de-identified prospective pilot data from Peking University First Hospital are not publicly available because of institutional ethics, privacy, and data governance restrictions. Individual participant data from the prospective pilot will not be shared publicly. Requests for aggregate results, analysis details, or additional methodological information can be directed to the corresponding authors after publication and will be considered subject to institutional approval and applicable data-use agreements.

### Code Availability

The code used for model development and evaluation in this study is publicly available at https://github.com/Tangoz1003/PocketED-K.

## Acknowledgements

Shenda Hong is supported by the National Natural Science Foundation of China (62102008), CCF-Tencent Rhino-Bird Open Research Fund (CCF-Tencent RAGR20250108), CCF-Zhipu Large Model Innovation Fund (CCF-Zhipu202414), PKU-OPPO Fund (BO202301, BO202503), Research Project of Peking University in the State Key Laboratory of Vascular Homeostasis and Remodeling (2025-SKLVHR-YCTS-02), and the Beijing Municipal Science and Technology Commission (Z251100000725008).

Xiaojing Li is supported by the Integrated Traditional Chinese and Western Medicine Chronic Disease Management Research Project (CXZH2024100).

The authors thank all collaborators and participating institutions for their support and contributions to this research.

## Author Contributions

Gongzheng Tang, Xiaojing Li contributed equally to this work. Gongzheng Tang contributed to methodology development, model implementation, validation, formal analysis, and drafting of the manuscript. Xiaojing Li contributed to result interpretation and manuscript revision. Yujie Xiao, Kexin Wang, Meng Xu, Xingliang Wu, Zheng Wei, Miao Yu, Xiaolan Chen, Wei Hong, Fang Cheng and Xiuqing Li contributed to data preparation and investigation. Xiaojing Li and Shenda Hong conceived and supervised the study, provided resources, guided study design and interpretation, and revised the manuscript. All authors reviewed and approved the final manuscript. Xiaojing Li and Shenda Hong are corresponding authors and take responsibility for the integrity of the work.

## References

1. Weiner, I.D., and Wingo, C.S. (1997). Hypokalemia–consequences, causes, and correction. Journal of the American Society of Nephrology 8, 1179–1188.

2. Jensen, H.K., Brabrand, M., Vinholt, P.J., Hallas, J., and Lassen, A.T. (2015). Hypokalemia in acute medical patients: risk factors and prognosis. The American journal of medicine 128, 60–67.

3. Coca, S.G., Perazella, M.A., and Buller, G.K. (2005). The cardiovascular implications of hypokalemia. American journal of kidney diseases 45, 233–247.

4. Gennari, F.J. (2002). Disorders of potassium homeostasis: hypokalemia and hyperkalemia. Critical care clinics 18, 273–288.

5. Cohn, J.N., Kowey, P.R., Whelton, P.K., and Prisant, L.M. (2000). New guidelines for potassium replacement in clinical practice: a contemporary review by the national council on potassium in clinical practice. Archives of internal medicine 160, 2429–2436.

6. Kovesdy, C.P., Matsushita, K., Sang, Y., Brunskill, N.J., Carrero, J.J., Chodick, G., Hasegawa, T., Heerspink, H.L., Hirayama, A., Landman, G.W. et al. (2018). Serum potassium and adverse outcomes across the range of kidney function: a ckd prognosis consortium meta-analysis. European heart journal 39, 1535–1542.

7. Goyal, A., Spertus, J.A., Gosch, K., Venkitachalam, L., Jones, P.G., Van den Berghe, G., and Kosiborod, M. (2012). Serum potassium levels and mortality in acute myocardial infarction. Jama 307, 157–164.

8. Helfant, R.H. (1986). Hypokalemia and arrhythmias. The American journal of medicine 80, 13–22.

9. Weiss, J.N., Qu, Z., and Shivkumar, K. (2017). Electrophysiology of hypokalemia and hyperkalemia. Circulation: arrhythmia and electrophysiology 10, e004667.

10. Palmer, B.F. (2010). A physiologic-based approach to the evaluation of a patient with hypokalemia. American journal of kidney diseases 56, 1184–1190.

11. Zhao, Q., Geng, S., Wang, B., Sun, Y., Nie, W., Bai, B., Yu, C., Zhang, F., Tang, G., Zhang, D. et al. (2024). Deep learning in heart sound analysis: from techniques to clinical applications. Health Data Science 4, 0182.

12. Luo, Y., Liu, X.Y., Yang, K., Huang, K., Hong, M., Zhang, J., Wu, Y., and Nie, Z. (2024). Toward unified ai drug discovery with multimodal knowledge. Health Data Science 4, 0113.

13. Zhang, S., Mu, W., Dong, D., Wei, J., Fang, M., Shao, L., Zhou, Y., He, B., Zhang, S., Liu, Z. et al. (2023). The applications of artificial intelligence in digestive system neoplasms: a review. Health Data Science 3, 0005.

14. Liu, X., Gao, K., Liu, B., Pan, C., Liang, K., Yan, L., Ma, J., He, F., Zhang, S., Pan, S. et al. (2021). Advances in deep learning-based medical image analysis. Health Data Science 2021, 8786793.

15. Xiao, Y., Tang, G., Liu, W., Li, J., Nie, G., Kan, Z., Zhang, D., Zhao, Q., and Hong, S. (2025). Anyecg-lab: An exploration study of fine-tuning an ecg foundation model to estimate laboratory values from single-lead ecg signals. arXiv preprint arXiv:2510.22301.

16. Nie, G., Zhu, J., Tang, G., Zhang, D., Geng, S., Zhao, Q., and Hong, S. (2024). A review of deep learning methods for photoplethysmography data. arXiv preprint arXiv:2401.12783.

17. Huang, S., Zhang, D., Fan, S., Tang, G., Geng, S., Xiao, Y., Wu, X., Yan, M., Wang, H., Zhang, R. et al. (2025). Opportunistic screening of wolff-parkinson-white syndrome using single-lead ai-ecg mobile system: A real-world study of over 3.5 million ecg recordings in china. arXiv preprint arXiv:2510.24750.

18. Poterucha, T.J., Jing, L., Ricart, R.P., Adjei-Mosi, M., Finer, J., Hartzel, D., Kelsey, C., Long, A., Rocha, D., Ruhl, J.A. et al. (2025). Detecting structural heart disease from electrocardiograms using ai. Nature 644, 221–230.

19. Rawlani, M., Ieki, H., Binder, C., Yuan, V., Chiu, I.M., Bhatt, A., Ebinger, J.E., Sahashi, Y., Ambrosy, A.P., Usuku, H. et al. (2025). Artificial intelligence prediction of age from echocardiography as a marker for cardiovascular disease. npj Digital Medicine 8, 688.

20. Nie, G., Zhao, Q., Tang, G., Li, Y., and Hong, S. (2025). Artificial intelligence-derived photoplethysmography age as a digital biomarker for cardiovascular health. Communications Medicine 5, 481.

21. Xiao, Y., Tang, G., Zhang, D., Li, J., Nie, G., Wang, H., Huang, S., Liu, T., Zhao, Q., Chen, K. et al. (2025). Fine-tuning an ecg foundation model to predict coronary ct angiography outcomes. arXiv preprint arXiv:2512.05136.

22. Oikonomou, E.K., Vaid, A., Holste, G., Coppi, A., McNamara, R.L., Baloescu, C., Krumholz, H.M., Wang, Z., Apakama, D.J., Nadkarni, G.N. et al. (2025). Artificial intelligence-guided detection of under-recognised cardiomyopathies on point-of-care cardiac ultrasonography: a multicentre study. The Lancet Digital Health 7, e113–e123.

23. Tang, G., Zhao, Q., Nie, G., Xiao, Y., Geng, S., Xie, D., Huang, S., Zhang, D., Yao, X., Wang, J., Chen, K., Zhang, L., and Hong, S. (2026). Artificial intelligence-enabled single-lead ecg for non-invasive hyperkalemia detection: development, multicenter validation, and proof-of-concept deployment. arXiv preprint arXiv:2603.14177.

24. Lin, C.S., Lin, C., Fang, W.H., Hsu, C.J., Chen, S.J., Huang, K.H., Lin, W.S., Tsai, C.S., Kuo, C.C., Chau, T. et al. (2020). A deep-learning algorithm (ecg12net) for detecting hypokalemia and hyperkalemia by electrocardiography: algorithm development. JMIR medical informatics 8, e15931.

25. Lin, C., Chau, T., Lin, C.S., Shang, H.S., Fang, W.H., Lee, D.J., Lee, C.C., Tsai, S.H., Wang, C.H., and Lin, S.H. (2022). Point-of-care artificial intelligence-enabled ecg for dyskalemia: a retrospective cohort analysis for accuracy and outcome prediction. NPJ digital medicine 5, 8.

26. Wang, C.X., Zhang, Y.C., Kong, Q.L., Wu, Z.X., Yang, P.P., Zhu, C.H., Chen, S.L., Wu, T., Wu, Q.H., and Chen, Q. (2021). Development and validation of a deep learning model to screen hypokalemia from electrocardiogram in emergency patients. Chinese medical journal 134, 2333–2339.

27. Collins, G.S., Moons, K.G., Dhiman, P., Riley, R.D., Beam, A.L., Van Calster, B., Ghassemi, M., Liu, X., Reitsma, J.B., Van Smeden, M. et al. (2024). Tripod+ ai statement: updated guidance for reporting clinical prediction models that use regression or machine learning methods. bmj 385.

28. Li, J., Aguirre, A.D., Junior, V.M., Jin, J., Liu, C., Zhong, L., Sun, C., Clifford, G., Brandon Westover, M., and Hong, S. (2025). An electrocardiogram foundation model built on over 10 million recordings. Nejm ai 2, AIoa2401033.

29. Radosavovic, I., Kosaraju, R.P., Girshick, R., He, K., and Dollár, P. (2020). Designing network design spaces. In Proceedings of the IEEE/CVF conference on computer vision and pattern recognition. pp. 10428–10436.

30. Hong, S., Xu, Y., Khare, A., Priambada, S., Maher, K., Aljiffry, A., Sun, J., and Tumanov, A. (2020). Holmes: health online model ensemble serving for deep learning models in intensive care units. In Proceedings of the 26th ACM SIGKDD International Conference on Knowledge Discovery & Data Mining. pp. 1614–1624.

